# Global and Regional Burden of bloodstream infection caused by *Candida spp.* from 1990-2021: A Systematic Analysis and Future Projections

**DOI:** 10.1101/2025.11.23.25340841

**Authors:** Hanli Wang, Min Xing, Haoyu Ji, Yusheng Cheng, Shuhui Hu, Guofeng Wang, Susheng Zhou, Yonghui Han, Chenghui Wang, Lei Zha, Qinghai You

## Abstract

**Purpose:** This study aimed to assess and predict the global and regional burden of candidemia.

**Methods:** We extracted data on the estimated burden of candidemia from 1990 to 2021 from the MICROBE database. The number of deaths, age-standardized mortality of rate(ASMR) and disability-adjusted life-years(DALYs) associated with candidemia, were systematically analyzed. The burden of candidemia over the next 30 years was predicted using the Bayesian age-period-cohort model with integrated nested Laplace approximations.

**Results:** In 2021, candidemia accounted for approximately 174,716 deaths(95% UI, 159,876–189,556) globally, with an ASMR of 2.28 (95% UI, 2.06–2.50) per 100k population. The highest burden of candidemia was observed in Southern, Western, and Eastern Sub-Saharan Africa, while the lowest burden occurred in High-income Asia Pacific, Australasia, and Western Europe. Males and older adults exhibited a higher disease burden, while the under-5 age group also demonstrated a substantial burden. Projections indicated that the mortality rate of candidemia was expected to decline to 1.72 per 100k(95% UI: 1.45–2.00) by 2050.

**Conclusions:** Our findings underscore the significant global burden of candidemia, with notable regional disparities. Addressing this burden requires enhanced surveillance, robust antifungal stewardship programs, targeted interventions for high-risk populations, and improved diagnostic and therapeutic capabilities to mitigate the impact of antifungal resistance and reduce mortality.

## Introduction

Candidemia, a life-threatening fungal infection caused by the hematogenous dissemination of *Candida species*, is defined by ≥1 positive blood culture and recognized as a leading cause of healthcare-associated bloodstream infections.[1] The condition is associated with diverse comorbidities and risk factors including prolonged hospitalization, parenteral nutrition, indwelling central venous catheters, broad-spectrum antibiotic exposure, glucocorticoid use, and major abdominal surgery.[2, 3] Candidemia frequently precipitates serious complications such as infective endocarditis, septic shock, endogenous endophthalmitis, osteomyelitis, and hepatic or splenic abscesses, substantially compromising patient prognosis and quality of life.[4] Notably, COVID-19-associated *Candida* infections have surged in recent years, further elevating mortality risks.[5] The attributable mortality ranges from 30–50%, reaching 77.3% in high-risk cohorts.[6, 7]

This complex clinical profile imposes significant economic burdens. A 2018 survey in America attributed $6.7 billion annually to fungal infections, with candidemia patients incurring >2-fold higher costs, prolonging hospital stays and mortality versus non-fungal cases. The economic burden is substantial, with inpatient costs in America averaging $40,424 per candidemia case and totaling $5.6 billion annually.[8] Candidemia typically presents as a fever or bloodstream infection that cannot be clinically distinguished from bacteremia without appropriate diagnostics. Although blood culture remains the gold standard, its limited sensitivity(approximately 50%) and delayed results impede early intervention[4]. Advanced diagnostics, including 1,3-β-D-glucan testing, Candida PCR, T2Candida®, and MALDI-TOF, show promise but also increase expenditures.[9] The escalating burden of candidemia underscores its status as an urgent public health priority. Nevertheless, evidence-based mitigation strategies, such as prophylactic antifungal therapy in high-risk populations including premature neonates, may reduce fungal colonization and invasive infections.[10] Crucially, timely implementation of core interventions including early diagnosis, rigorous source control, prompt appropriate antifungal therapy collectively reduced mortality[4]. The persistence of high clinical and economic burden despite therapeutic advances highlights the critical need for comprehensive, evidence-based prevention and control programs.

Global surveillance programs revealed significant epidemiological trends in candidemia. According to CDC’s Emerging Infections Program (EIP) surveillance data, the crude incidence reached 8.7 cases per 100,000(100k) population (n=3,492 cases), with substantial variations across regions, age groups, sexes, and racial groups.[2] The all-cause in-hospital mortality rate for candidemia was as high as 25%, with an 8% mortality rate within 48 hours of diagnosis. The European Confederation of Medical Mycology Candida III study furthered highlighted the significant impact of candidemia on hospital stays, particularly in ICU, thus intensifying the burden on healthcare systems. This study reported an overall mortality rate of 40.4%, with attributable mortality varying by the causative Candida species. For example, *Candida tropicalis* infections were associated with an attributable mortality rate of 63.6%. Additionally, candidemia was found to significantly extend hospital stays, which, especially in ICU settings, further strains healthcare resources.[11]

Recent studies had utilized data from various countries to assess the burden of candidemia;[1, 11] however, a comprehensive estimation of the overall burden and its correlation with socioeconomic development remained unexplored. In this study, we conducted a systematic analysis of the estimated burden of candidemia across 21 Global Burden of Disease (GBD) regions from 1990 to 2021, using data from the MICROBE (Measuring Infectious Causes and Resistance Outcomes for Burden Estimation) project. Furthermore, analyzing global temporal trends and projecting future trajectories of candidemia is essential, as it provides valuable insights for developing targeted prevention and control strategies.

## Material and methods

### Overview data sources

This study utilized data from the MICROBE database, an open-access repository developed by the Institute for Health Metrics and Evaluation (IHME). MICROBE provides extensive global estimates of the burden of pathogens, including candidemia, across 204 countries and territories, 21 Global Burden of Disease (GBD) regions, 7 super-regions, and global aggregates from 1990 to 2021. The dataset includes data on mortality, disability-adjusted life years (DALYs), age-standardized mortality of rate(ASMR) and age-standardized DALYs of rate(ASDR), which are derived from systematic reviews, surveillance data, and modeling frameworks. The methodologies for data collection, quality adjustments, and uncertainty quantification were fully detailed in previous GBD studies[12]. All data used in this study were extracted directly from the MICROBE platform(accessible at: https://vizhub.healthdata.org/microbe/).

### Statistical Analysis

ASMR and DALYs were computed using the GBD reference population, allowing for comparisons across regions and over time. To examine trends from 1990 to 2021, we calculated the average annual percentage change (AAPC) in age-standardized rates using joinpoint regression. AAPC values, along with their 95% uncertainty intervals (UIs), were calculated globally and for each GBD region, broken down by sex and age group. Sociodemographic Index(SDI) is a composite index that incorporates three key indicators including income, education, and fertility rate that together reflect the socio-economic development of a population. The SDI scale ranges from 0 to 1, with higher values indicating greater development. In this study, SDI was used to explore its association with the burden of candidemia-related mortality and DALYs, both globally and across the 21 GBD regions.

The Age-Period-Cohort (APC) model, proposed by Mason in 1973, is an epidemiological tool used to assess the impact of age, period, and cohort effects on disease outcomes. This model decomposes the effects into three components: age effects, which reflect developmental changes and accumulated exposures; period effects, which represent temporal changes affecting all age groups; and cohort effects, which capture differences between groups born in the same time period. The APC model is expressed as a multiple regression, where the outcome is a function of these three factors. To improve the robustness of our projections, we applied the Bayesian Age-Period-Cohort (BAPC) model with integrated nested Laplace approximations (INLA) to forecast candidemia burden from 2022 to 2050.[13] The BAPC model builds on the APC framework by incorporating Bayesian inference with INLA. This approach enhances precision and coverage over traditional APC models by smoothing the priors for age, period, and cohort effects using a second-order random walk. It also avoids the challenges of Markov Chain Monte Carlo sampling, providing more reliable posterior predictions, particularly for long-term disease burden trends. All analyses were conducted using publicly available aggregate data, and all computations were carried out in R software (version 3.62).

## Results

### Epidemiological profile of candidemia in 2021

In 2021, candidemia caused 174,716 deaths globally, reflecting a 29.7% increase in absolute mortality compared to 1990. However, ASMR decreased by 19.7%, from 2.84 to 2.28 per 100k population, indicating progress in healthcare despite population growth and aging. Similarly, the global DALYs attributed to candidemia decreased to 7,424,411, with ASDR dropping from 140 to 106 per 100k. Notably, males consistently showed higher mortality and DALYs across most age groups compared to females, except among individuals aged 80 years and older, where female mortality exceeded male mortality, likely due to longer life expectancy(Table 1, S1).

**Table 1.** Mortalitys in number and age standardized rates per 100k of Candidemia in 1990, 2019, 2021, by Global Burden of Disease region(generated from data available at VizHub - MICROBE (healthdata.org)).

|  | Deaths in 1990 |  | Deaths in 2019 |  | Deaths in 2021 |  | AAPC (95% UI) |
| --- | --- | --- | --- | --- | --- | --- | --- |
|  | Number (95% UI) | ASRs per 100k(95%UI) | Number (95% UI) | ASRs per 100k(95%UI) | Number (95% UI) | ASRs per 100k(95%UI) |  |
| Global | 134698(120850 to 148547) | 2.84(2.60 to 3.09) | 174927(158179 to 191675) | 2.34(2.10 to 2.58) | 174716(159876 to 189556) | 2.28(2.06 to 2.50) | -0.71(-0.78 to -0.64) |
| Female | 62839(56784 to 68894) | 2.56(2.34 to 2.79) | 81261(73079 to 89443) | 2.07(1.85 to 2.29) | 81126(73887 to 88364) | 2.01(1.81 to 2.20) | -0.79(-0.87 to -0.71) |
| Male | 71859(63805 to 79914) | 3.17(2.89 to 3.45) | 93666(84111 to 103222) | 2.66(2.38 to 2.94) | 93590(85104 to 102077) | 2.60(2.35 to 2.85) | -0.63(-0.71 to -0.55) |
| Central Europe Eastern Europe | 12901(11930 to 13873) | 3.00(2.76 to 3.24) | 14300(13434 to 15167) | 2.50(2.36 to 2.65) | 14156(13073 to 15240) | 2.44(2.26 to 2.61) | -0.64(-0.84 to -0.43) |
| Central Asia | 1544(1353 to 1736) | 2.62(2.37 to 2.88) | 1860(1699 to 2020) | 2.32(2.13 to 2.50) | 1840(1661 to 2019) | 2.23(2.02 to 2.43) | -0.47(-0.80 to -0.15) |
| Central Europe | 4021(3756 to 4286) | 3.04(2.83 to 3.25) | 4331(4046 to 4615) | 2.12(1.99 to 2.24) | 4314(3940 to 4688) | 2.07(1.90 to 2.24) | -1.22(-1.42 to -1.02) |
| Eastern Europe | 7336(6749 to 7922) | 3.00(2.76 to 3.25) | 8103(7565 to 8641) | 2.62(2.46 to 2.77) | 7996(7228 to 8764) | 2.55(2.33 to 2.78) | -0.47(-0.98 to 0.04) |
| High-income | 23666(22112 to 25220) | 2.14(2.01 to 2.27) | 31281(28013 to 34550) | 1.56(1.43 to 1.69) | 31318(27964 to 34672) | 1.50(1.38 to 1.62) | -1.16(-1.28 to -1.03) |
| Australasia | 442(414 to 470) | 1.99(1.87 to 2.12) | 704(631 to 776) | 1.45(1.32 to 1.58) | 714(639 to 789) | 1.37(1.25 to 1.50) | -1.18(-1.34 to -1.03) |
| High-income Asia Pacific | 2854(2665 to 3044) | 1.61(1.50 to 1.73) | 4766(4104 to 5428) | 1.08(0.97 to 1.18) | 4928(4209 to 5648) | 1.05(0.94 to 1.16) | -1.42(-1.49 to -1.36) |
| High-income North America | 6762(6322 to 7201) | 2.02(1.90 to 2.14) | 10401(9432 to 11370) | 1.80(1.66 to 1.94) | 10662(9685 to 11640) | 1.77(1.64 to 1.91) | -0.43(-0.55 to -0.30) |
| Southern Latin America | 1182(1103 to 1262) | 2.54(2.38 to 2.71) | 1648(1538 to 1759) | 2.13(1.99 to 2.27) | 1539(1429 to 1650) | 1.93(1.79 to 2.06) | -0.88(-1.20 to -0.56) |
| Western Europe | 12426(11534 to 13318) | 2.33(2.18 to 2.48) | 13763(12266 to 15259) | 1.53(1.40 to 1.66) | 13474(11947 to 15001) | 1.45(1.32 to 1.57) | -1.53(-1.68 to -1.38) |
| Latin America and Caribbean | 8238(7366 to 9109) | 2.48(2.27 to 2.68) | 11072(10026 to 12117) | 2.01(1.81 to 2.22) | 11006(9837 to 12175) | 1.93(1.72 to 2.15) | -0.81(-0.95 to -0.66) |
| Andean Latin America | 1040(912 to 1168) | 2.69(2.42 to 2.96) | 1243(1049 to 1437) | 2.13(1.80 to 2.46) | 1135(923 to 1347) | 1.91(1.56 to 2.27) | -1.08(-1.20 to -0.96) |
| Caribbean | 796.81(703.95 to 889.67) | 2.44(2.20 to 2.68) | 1091(960 to 1221) | 2.33(2.03 to 2.63) | 1097(936 to 1259) | 2.30(1.94 to 2.65) | -0.18(-0.38 to 0.03) |
| Central Latin America | 3342(2953 to 3730) | 2.36(2.16 to 2.56) | 4687(4246 to 5128) | 2.07(1.86 to 2.28) | 4723(4133 to 5313) | 2.03(1.76 to 2.29) | -0.48(-0.66 to -0.31) |
| Tropical Latin America | 3058(2724 to 3392) | 2.57(2.34 to 2.80) | 4055(3704 to 4406) | 1.85(1.67 to 2.03) | 4055(3738 to 4372) | 1.76(1.61 to 1.91) | -1.20(-1.29 to -1.10) |
| North Africa and Middle East | 9280(7882 to 10677) | 2.88(2.57 to 3.18) | 9964(8811 to 11116) | 2.22(1.99 to 2.45) | 9632(8498 to 10766) | 2.12(1.89 to 2.35) | -0.98(-1.05 to -0.91) |
| South Asia | 30727(25467 to 35987) | 2.74(2.39 to 3.08) | 34193(29293 to 39093) | 2.40(2.07 to 2.73) | 33793(29569 to 38017) | 2.34(2.05 to 2.62) | -0.50(-0.68 to -0.32) |
| Southeast Asia East Asia Oceania | 30140(26736 to 33545) | 2.45(2.19 to 2.72) | 42643(36939 to 48348) | 1.93(1.68 to 2.18) | 43809(38077 to 49541) | 1.90(1.67 to 2.13) | -0.82(-1.02 to -0.63) |
| East Asia | 21552(18887 to 24218) | 2.55(2.23 to 2.86) | 31060(25843 to 36277) | 1.79(1.50 to 2.08) | 31963(26435 to 37490) | 1.75(1.47 to 2.02) | -1.27(-1.56 to -0.97) |
| Oceania | 106(86 to 126) | 2.22(1.85 to 2.60) | 216(172 to 261) | 2.18(1.82 to 2.53) | 226(182 to 269) | 2.16(1.83 to 2.50) | -0.11(-0.23 to 0.00) |
| Southeast Asia | 8480(7353 to 9607) | 2.23(1.99 to 2.46) | 11370(10070 to 12671) | 2.02(1.79 to 2.25) | 11624(10334 to 12913) | 1.99(1.77 to 2.21) | -0.38(-0.48 to -0.27) |
| Sub-Saharan Africa | 19750(16616 to 22884) | 3.26(2.87 to 3.65) | 31478(25357 to 37599) | 3.08(2.63 to 3.53) | 19750(16616 to 22884) | 3.26(2.87 to 3.65) | -0.28(-0.39 to -0.16) |
| Central Sub-Saharan Africa | 1905(1504 to 2306) | 3.02(2.58 to 3.46) | 3094(2308 to 3879) | 2.80(2.23 to 3.37) | 2967(2146 to 3789) | 2.73(2.17 to 3.30) | -0.30(-0.45 to -0.16) |
| Eastern Sub-Saharan Africa | 7976(6807 to 9146) | 3.31(2.93 to 3.69) | 10848(8646 to 13051) | 2.89(2.44 to 3.34) | 10634(8203 to 13065) | 2.83(2.38 to 3.29) | -0.50(-0.59 to -0.41) |
| Southern Sub-Saharan Africa | 1351(1154 to 1548) | 2.82(2.51 to 3.14) | 1993(1749 to 2236) | 3.21(2.87 to 3.56) | 2024(1750 to 2298) | 3.21(2.83 to 3.59) | 0.41(0.26 to 0.56) |
| Western Sub-Saharan Africa | 8518(7050 to 9986) | 3.36(2.90 to 3.81) | 15543(12414 to 18672) | 3.18(2.70 to 3.66) | 15380(11998 to 18762) | 3.10(2.58 to 3.62) | -0.25(-0.32 to -0.19) |

95% UI=95% uncertainty intervals; ASRs per 100k= age standardised rates per 100 000

Regionally, Sub-Saharan Africa remained the highest-burden region, with Western, Eastern, and Southern Africa reporting the highest ASMR for candidemia in 2021(3.21, 3.10, and 2.83 per 100k, respectively). In contrast, high-income Asia-Pacific, Australasia, and Western Europe (1.05, 1.37, 1.45 per 100k, respectively) recorded the lowest rates. These trends were consistent for both ASMR and ASDR.

On a national level, China and India collectively accounted for 31.24% of global candidemia-related deaths in 2021, each reporting over 10,000 deaths, primarily due to their large populations. The ASMR ranged from 0.61 to 3.86 per 100k across various countries. Small nations(Tokelau, Lesotho, South Sudan) faced disproportionately high ASMR, while Singapore, San Marino, and Japan had some of the lowest (Figure 1). In 2021, age-specific patterns revealed a bimodal distribution of mortality, with peaks in children under 5 years (7.78 per 100k in males and 6.34 per 100k in females) and adults over 70 years (44.24 per 100k in males and 43.97 per 100k in females) (Figure 2). Central Europe, Eastern Europe, and Central Asia had the highest mortality rates for older adults, at 16.27 per 100k, while South Asia reported the lowest rates for those aged 5–49 years, at 0.28 per 100k (Figure 3). For further details on DALYs, refered to Figure S2, S3.

**Figure 1.**
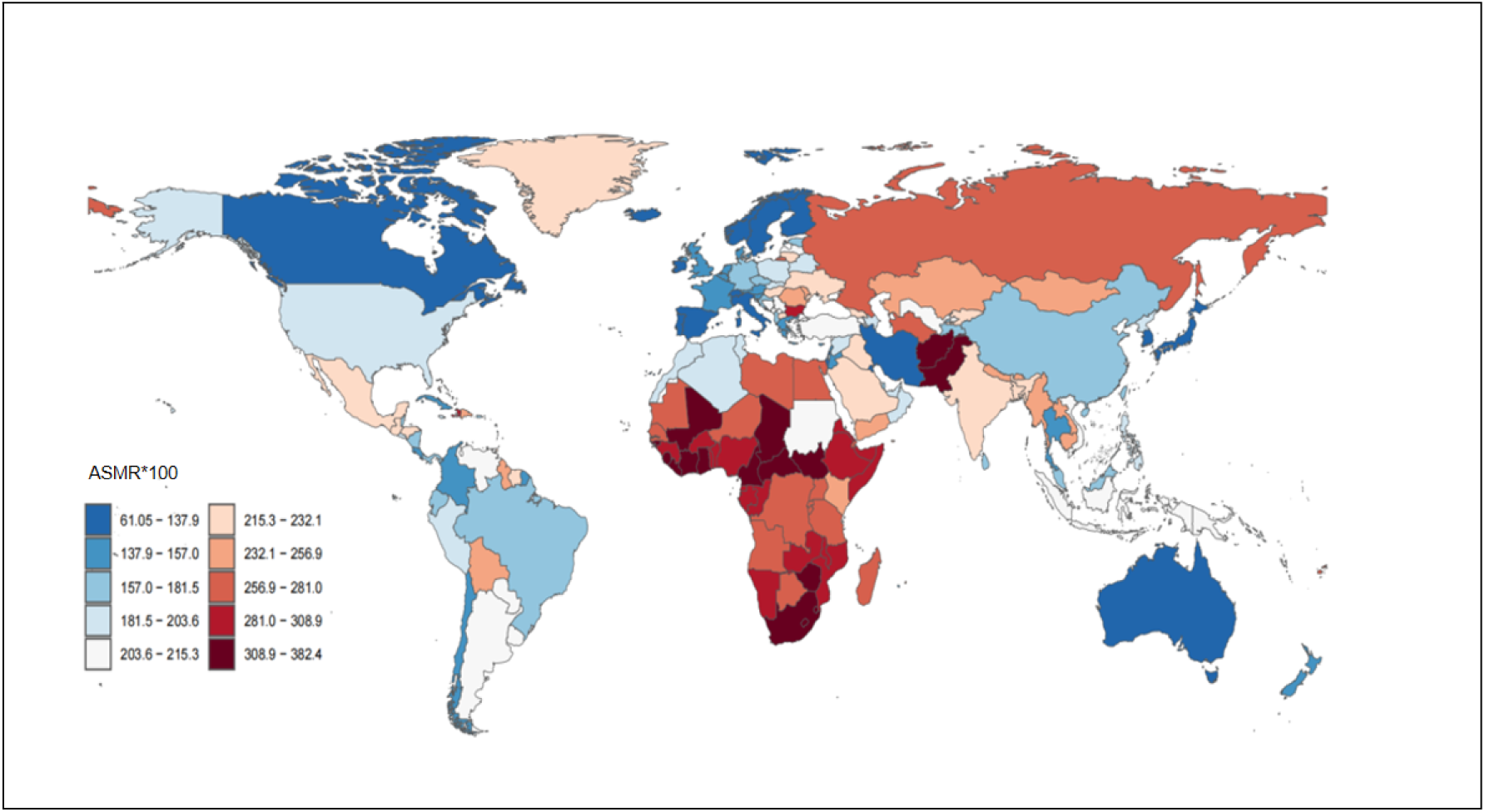
Age standardized mortality rates of Candidemia in the 204 countries and territories. The value in the figure is one hundred times that of ASMR.

**Figure 2.**
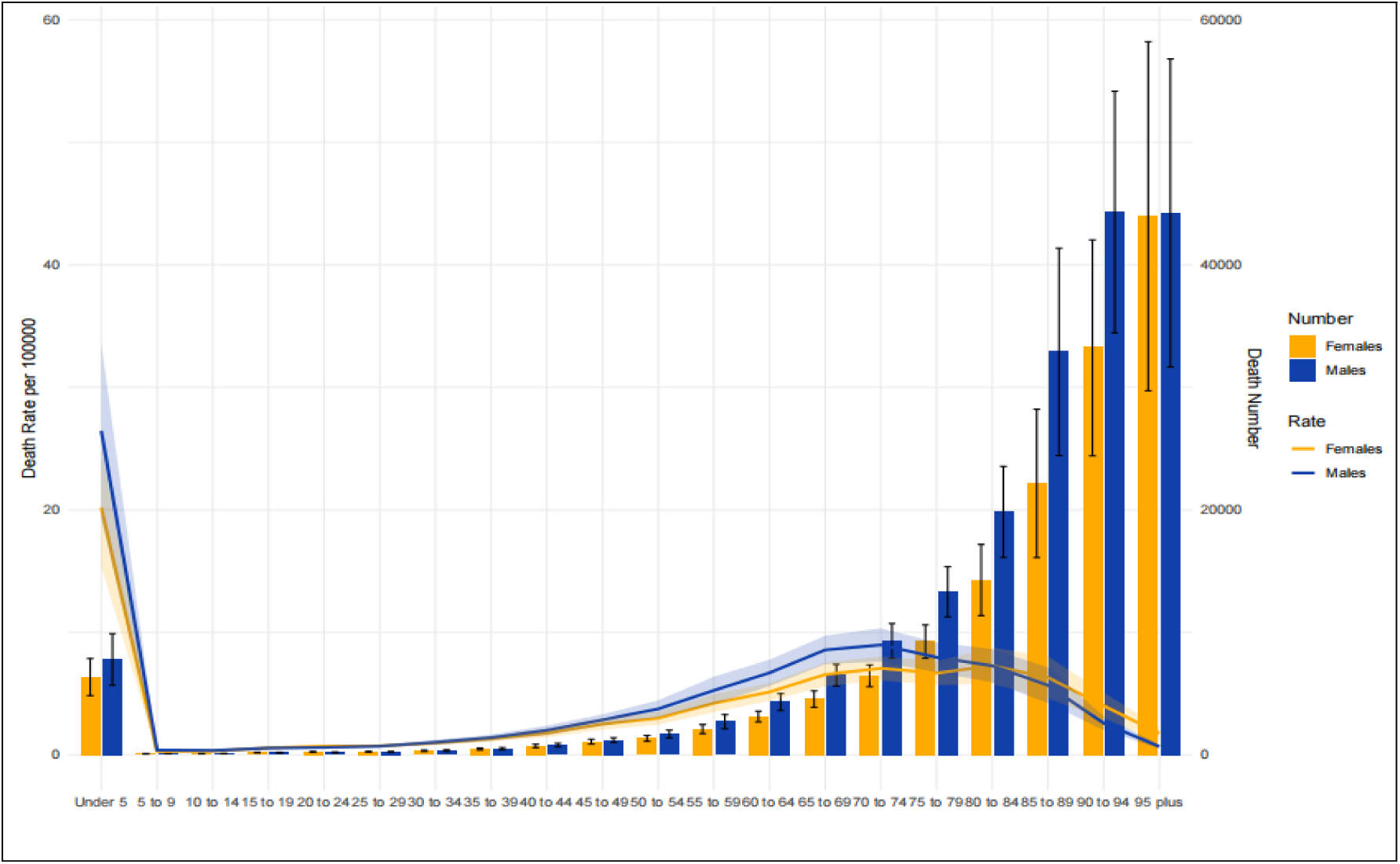
Mortalitys number and mortality rates between males and females with Candidemia across different age groups.

**Figure 3.**
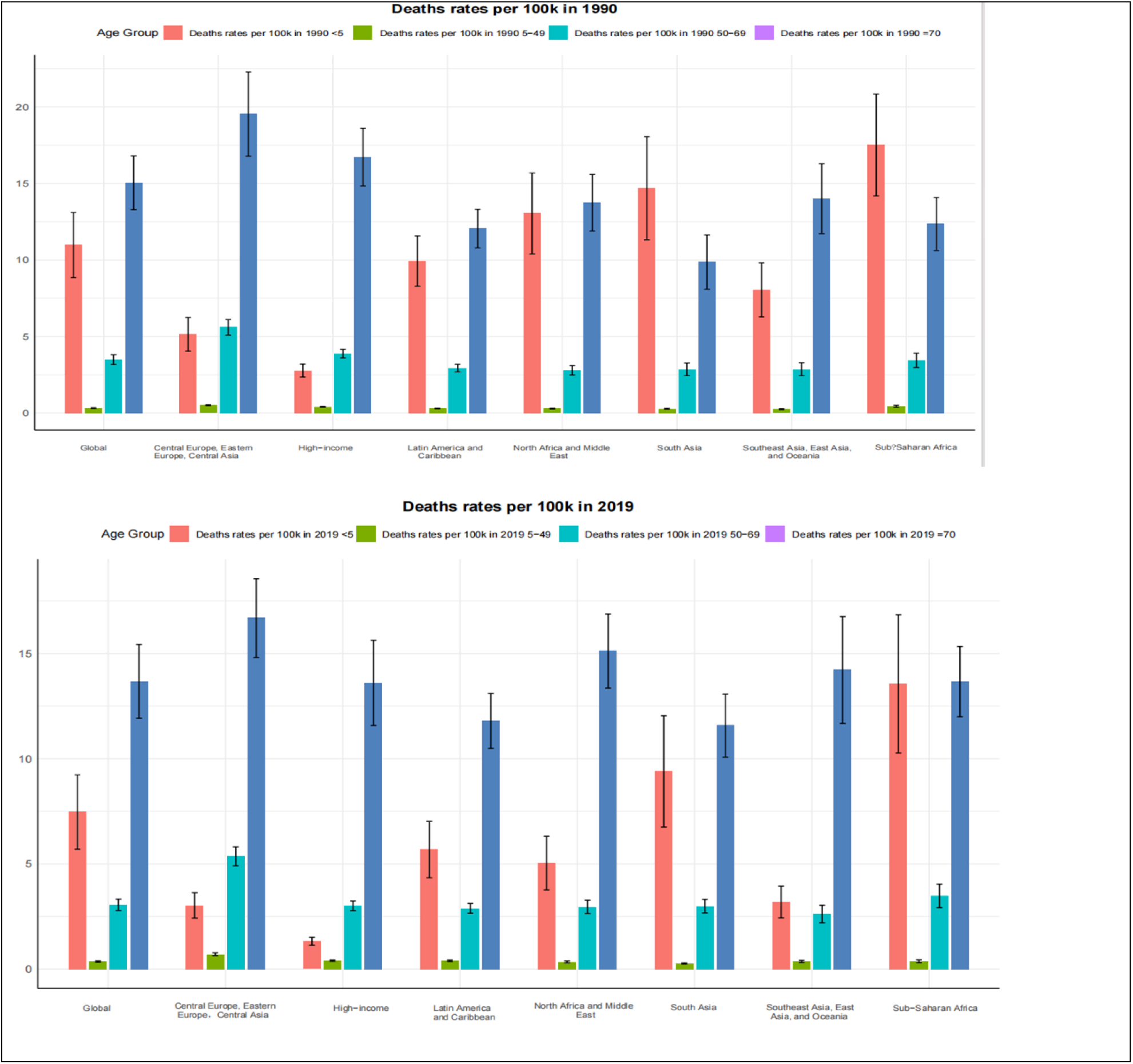
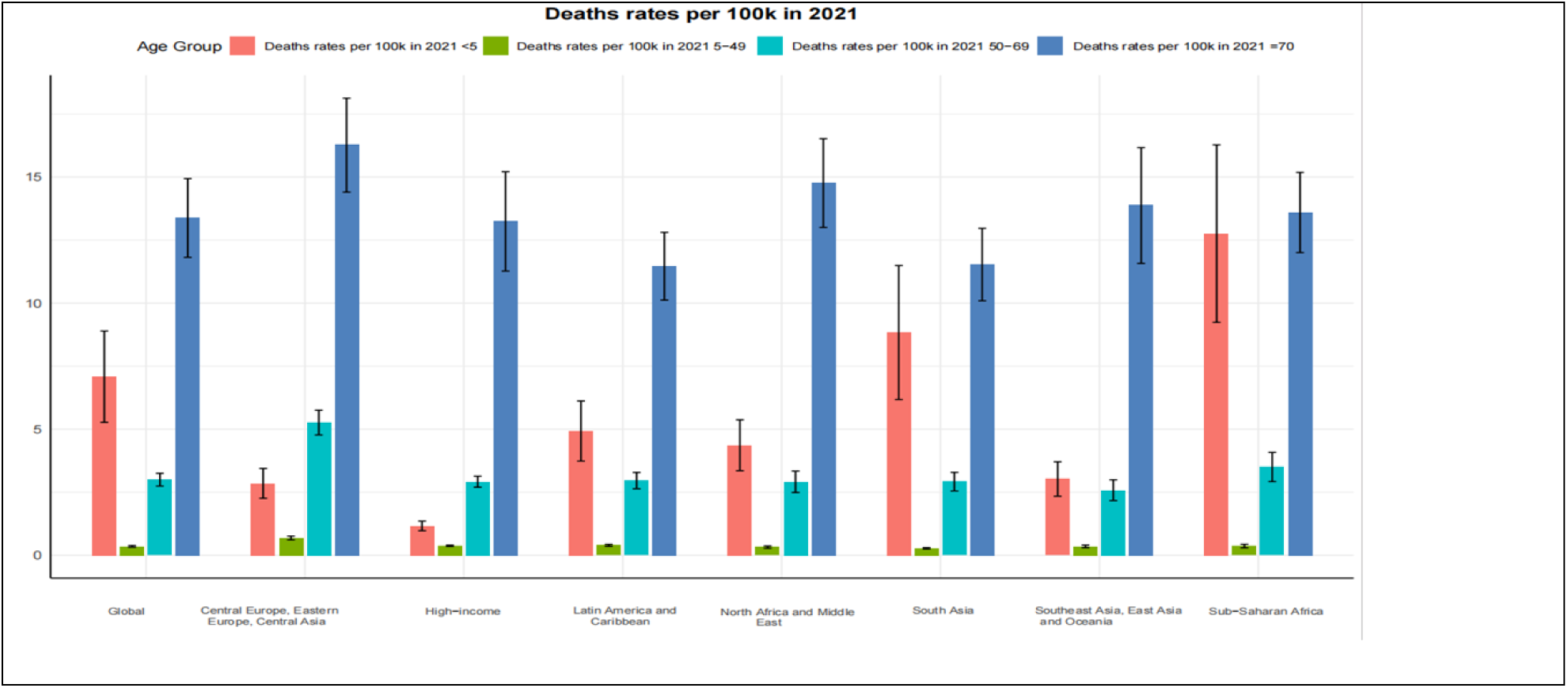
Number of total global and 7 super regional mortality with Candidemia in 1990, 2019, 2021.

### Temporal trends in candidemia mortality across different age group

Globally, ASMR for candidemia had decreased at an AAPC of -0.71%, with females experiencing a faster reduction(-0.79%) than males(-0.63%). This decline was driven by advancements in antifungal therapies, infection control, and neonatal care. However, despite these advancements, absolute mortality has risen, primarily due to population aging. Notably, the ≥70-year age group saw a dramatic increase in mortality by 117.6%, from 30,383 to 66,142 deaths, surpassing the mortality rate in the under-5 age group by 2015. Regionally, Western Europe (-1.53% AAPC), high-income Asia-Pacific (-1.42%), and East Asia (-1.27%) achieved the most substantial reductions in ASMR. In contrast, Sub-Saharan Africa experienced an increase in mortality (AAPC: 0.41%), driven by factors such as HIV or AIDS, limited healthcare access, and growing antifungal resistance. The greatest declines in ASDR were observed in East Asia (-2.28%) and Central Europe (-1.98%), while Oceania saw minimal change (-0.11%) (Table 1, Figure 4). National trends revealed stark disparities, with countries such as Tokelau, Lesotho, and Zimbabwe reporting the highest AAPC, indicating worsening mortality trends, whereas high-income nations like Singapore and Japan maintained low and stable mortality rates (Figure S6).

**Figure 4.**
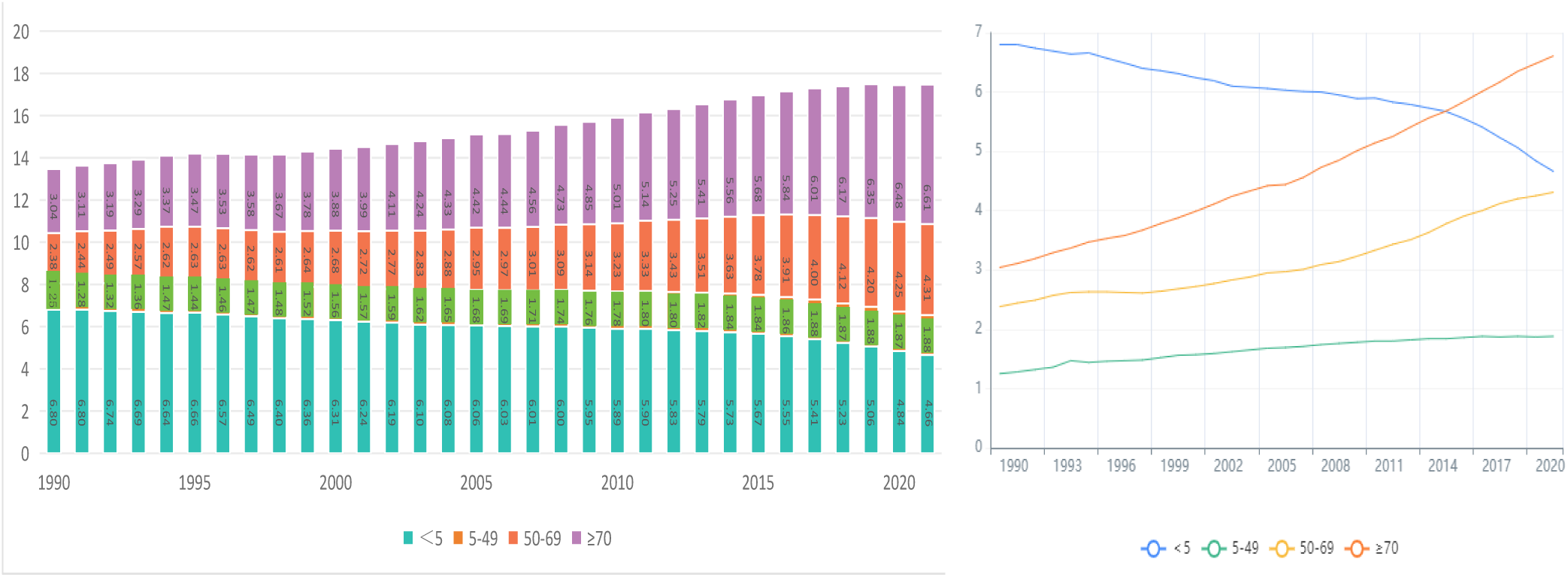
The mortality number of Candidemia(x 10, 000) from 1990-2021. Different colors represents different age.

### Association with sociodemographic index

At the regional level, a negative correlation was observed between SDI and both ASMR and ASDR of candidemia, from 1990-2021. As SDI increased, ASMR generally showed a slight decline, particularly up to an SDI value of approximately 0.7, after which the rate of decline accelerated more sharply. However, certain regions displayed unusual patterns. Western Sub-Saharan Africa, Central Europe, and Eastern Europe reported ASMR values higher than expected based on their SDI, while Oceania, Central Latin America, and Southeast Asia exhibited lower-than-expected ASMR values. Overall, most regions followed expected trends in line with their SDI, with the exception of Southern Sub-Saharan Africa, which displayed an inverted V-shaped correlation between SDI and ASMR (Figure 5, Figure S5).

**Figure 5.**
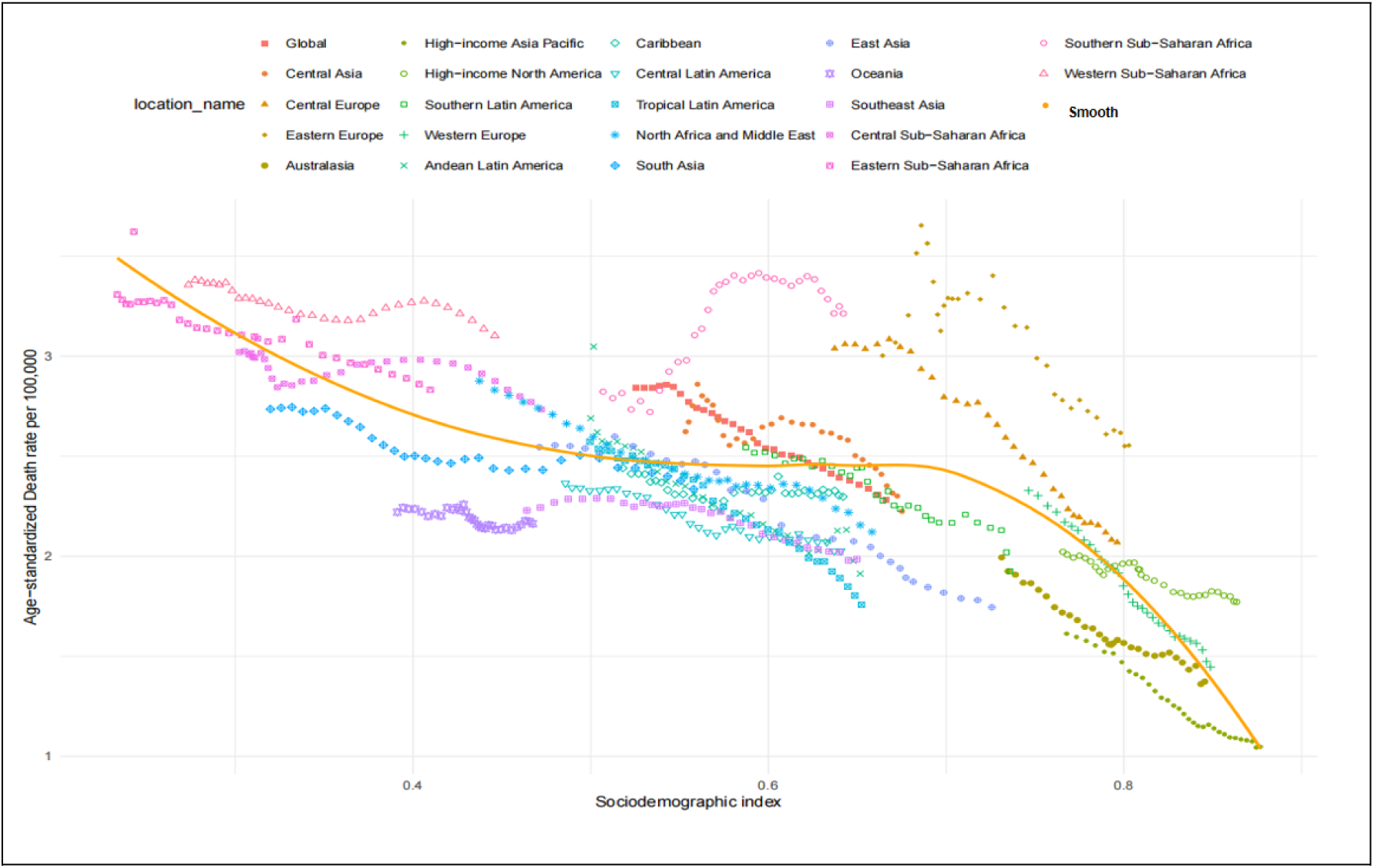
Age-standardized mortality rates of Candidemia for the 21 global burden of disease regions by sociodemographic index, 1990-2021. Thirty points are plotted for each region and exhibit the observed age-standardized mortality rates from 1990-2021 for that region. Expected values, based on SDI and mortality rates in all locations, are shown as a solid line. Regions above the solid line represent a higher than expected burden(eg, Western Sub−Saharan Africa) and regions below the line show a lower than expected burden(eg, Central Latin America).

### APC model

APC model revealed several notable trends. One key finding was that candidemia mortality rates increased with age across most age groups, except for the under-5 age group, where a declining trend was observed. Globally, mortality rates for candidemia generally declined across all age groups, reflecting improvements in overall mortality outcomes. The net drift in mortality rates was calculated to be -0.53%, indicating a modest reduction in candidemia related deaths over the study period. Local drift patterns also suggested that age variations had significant effects on mortality trends, with most age groups showing improvements in mortality rates. Age-related effects on candidemia mortality followed an expected exponential increase, with mortality rates rising steadily after the age of 5 and accelerating significantly after the age of 70. Both period and cohort effects mirrored these trends, indicating consistent improvements in candidemia related mortality across time periods and birth cohorts (Figure 6).

**Figure 6.**
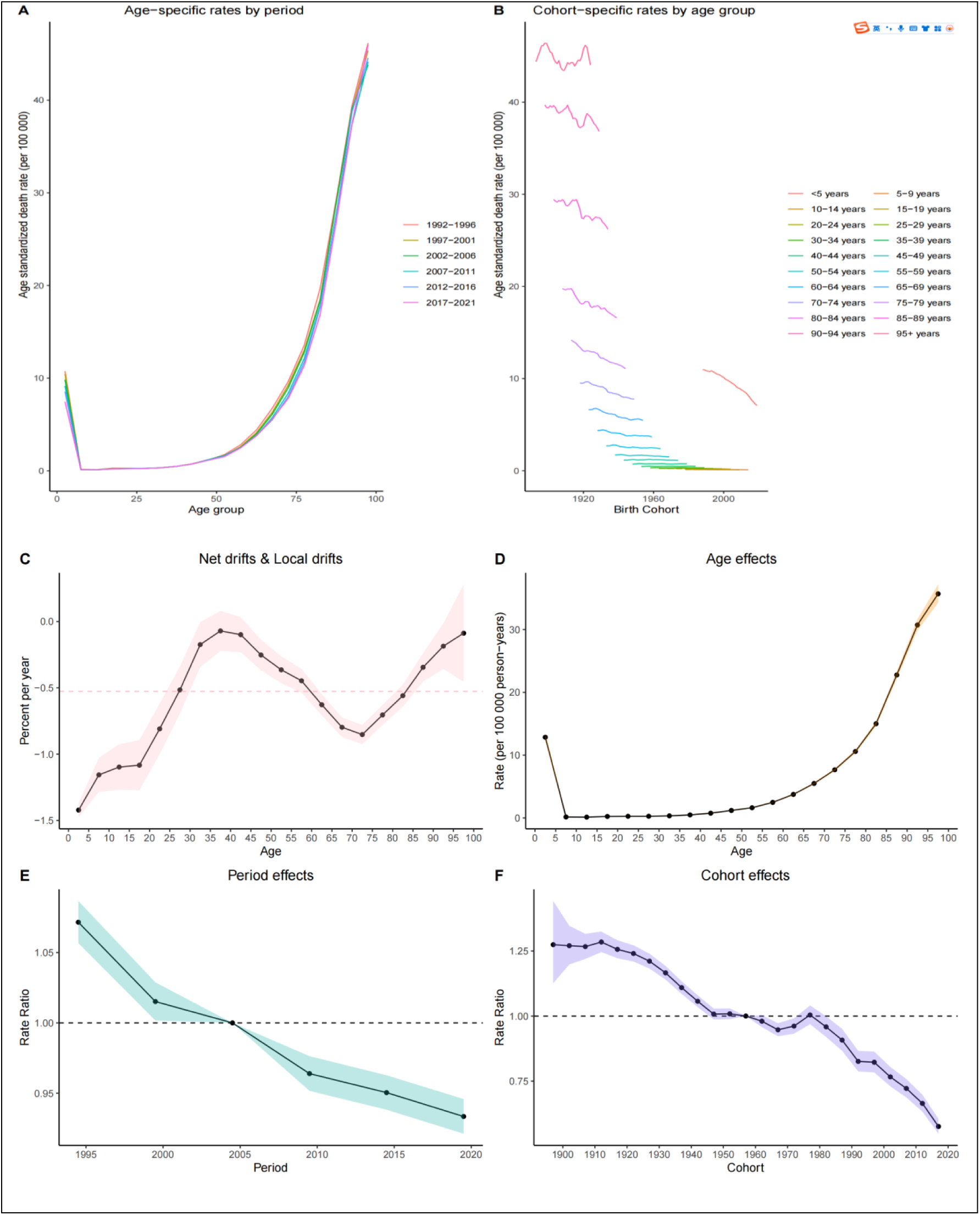
Age standardized mortality rates of Candidemia by age period cohort across global.(A) The age-specific mortality rates according to time periods; each line connects the age-specific mortality for a 5-year period. (B) The birth cohort-specific mortality rates according to age group; each line shows the birth cohort-specific mortality for 5-year age group. (C) Net drifts represents the overall annual percentage change and local drift values represent annual percentage change in each age group; The value of either one is less than 0 indicating a decreased trend in Candidemia mortality across the period. (D) Fitted longitudinal age curves of Candidemia mortality per 100k person-years. (E) Relative risk of each period adjusted for age and nonlinear cohort effects. (F) Relative risk of each cohort adjusted for age and nonlinear period effects.

Globally, ASMR for candidemia exhibited a persistent decline, mirroring trends observed across seven super-regions. However, substantial regional disparities are projected to persist through 2050. Projections indicate that ASMR will remain elevated in Central Europe, Eastern Europe, and Central Asia (2.10 per 100k), Sub-Saharan Africa (2.17 per 100k), and South Asia (1.75 per 100k) compared to the global average (1.72 per 100k), highlighting uneven progress in reducing candidemia related mortality (Figure 7).

**Figure 7.**
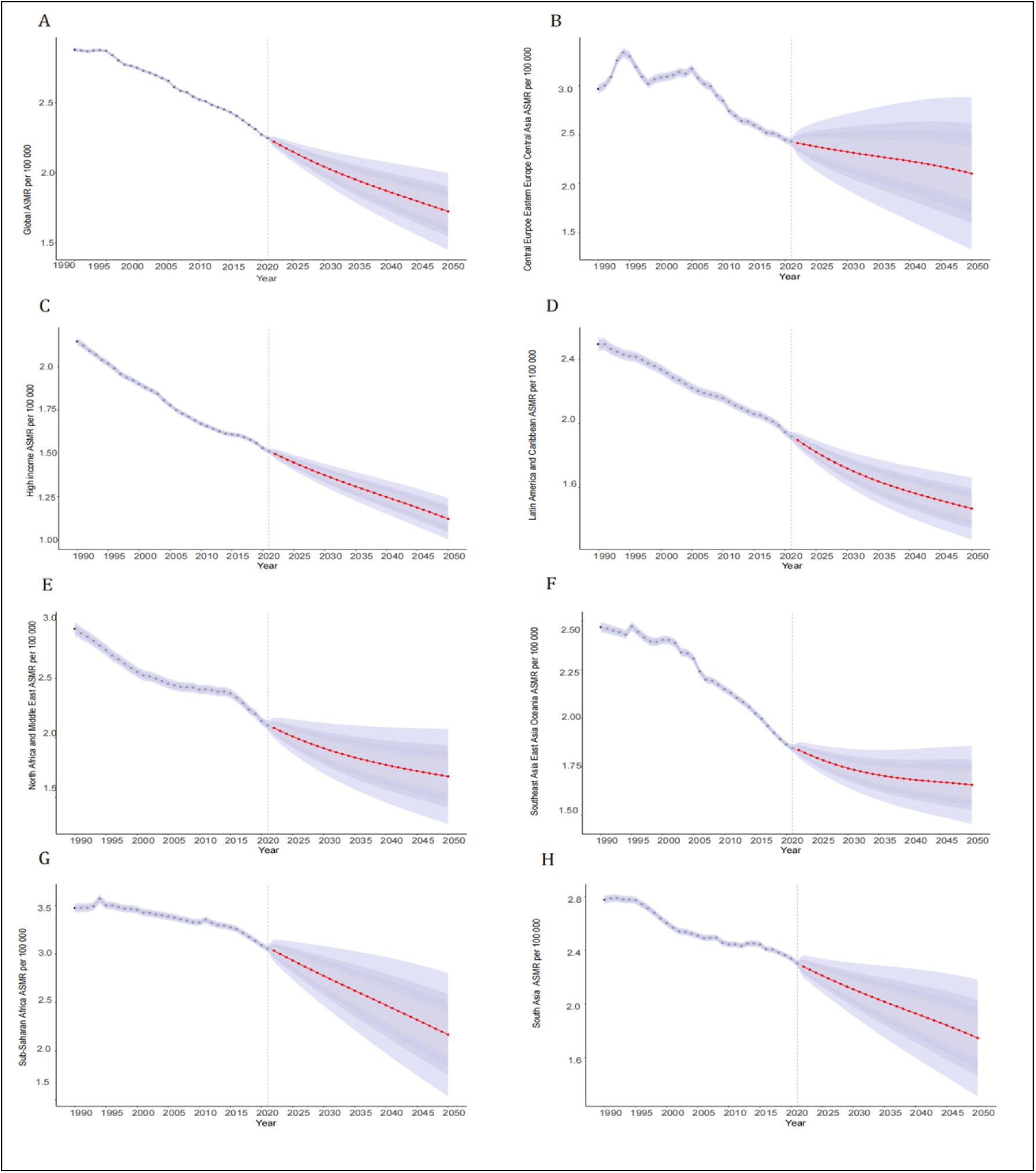
Trends in Candidemia-related age standardized mortality rates in total global and 7 super region(A-H): Observed(before 2021) and predicted(after 2021) age standardized mortality rates.

## Discussion

Global mortality due to candidemia had increased over the past few decades, rising from 134,698 cases in 1990 to 174,716 cases in 2021. Despite this rise in absolute mortality, ASMR had consistently declined, decreasing from 2.84 per 100k in 1990 to 2.28 per 100k in 2021. This decline in ASMR aligned with recent epidemiological studies, indicating a reduction in the relative global burden of candidemia globally.[11] Predictive model will forecast a further decline in the global ASMR to 1.72 per 100,000 by 2050, attributed to advancements in clinical diagnosis and treatment. This trend reflects significant progress in addressing candidemia, particularly in high-income countries. However, the concurrent increase in absolute mortality cases underscored that the global burden of candidemia remained substantial. This burden was disproportionately borne by resource-limited regions, such as Sub-Saharan Africa, primarily due to infrastructural deficits, resource constraints, multidimensional challenges in antifungal stewardship, insufficient healthcare facilities, and a higher underlying burden of predisposing conditions.[14]

Significant regional variations exist in candidemia burden and mortality. Mortality rates in high-income countries were 26% in Japan[15] and 21% in Australia[16]. In the United StatesAn estimated 5,628 deaths occurred among hospitalized candidemia patients, corresponding to a 25% mortality rate.[17] Infectious Disease Society of America, the CDC and National Healthcare Safety Network provided infection control guidance for healthcare institutions. The EQUAL scores was introduced by the European Confederation of Medical Mycology to measure the quality of candidemia management.[18] These guidelines emphasize the importance of hand hygiene, environmental disinfection, and infection prevention measures, including central venous catheters removal, early initiation of appropriate antifungal therapy, and prophylactic antifungal treatment.[14] However, the burden remains severe in low-income regions. Crude mortality rate reached 60% in Sub-Saharan Africa[19] and 56.7% for 30-day crude mortality in North Africa and the Middle East,[20] consistent with our findings. This elevated burden was associated with weak infrastructure, limited healthcare resources, antibiotic overuse, inadequate diagnostic tools, and insufficient antifungal treatment knowledge.[14] To sustainably reduce the global disease burden, increased healthcare investment in resource-limited regions, enhanced diagnostic capabilities, and reduced drug resistance are imperative. Concurrently, addressing regional disparities and high-risk group needs requires targeted public health resource allocation. African countries are addressing fungal disease burdens through regional collaborative networks, grassroots diagnostic capacity strengthening, and local expertise development.[21]

Candidemia burden is influenced by multiple factors. Consistent with prior research[17], our study found higher disease burden in males than females, suggesting greater challenges in infection management among males. This disparity may relate to gender differences in high-risk behaviors, comorbidities and healthcare access, highlighting the need for gender-specific interventions globally, especially in male-dense, resource-limited areas. Mortality burden also varied significantly by age, with markedly higher rates in children under 5 years and elderly populations, aligning with CDC’s EIP data showing elevated all-cause in-hospital mortality in these age groups.[2, 17] Contributing factors included comorbidities and immunosenescence in the elderly and immature immunity in young children. ICU admission, immunodeficiency, high Charlson Comorbidity Index, and infections by *Candida tropicalis* or *Candida auris* further increase mortality risk.[11, 18] Studies noted a shift in *Candida species* distribution consistent with SENTRY Antifungal Surveillance Program data.[22] Rising prevalence of fluconazole-resistant *non-albicans Candida species* including *Candida auris* and *Candida parapsilosis* may exacerbate economic burdens and mortality rates.

We observed no significant global or regional burden differences between 2019–2021, which contrasted with the findings of another study.[23, 24] CDC’s EIP data revealed that 25.5% of candidemia patients had COVID-19, which was associated with acute risk factors(ICU care, mechanical ventilation, central catheters, immunosuppressant use) and significantly higher in-hospital mortality(62.5% vs. 32.1%).[24] Another study reported 100% mortality in COVID-19-associated candidemia despite antifungal treatment, suggesting COVID-19 as a novel risk factor.[23] Our inclusion of non-ICU or non-COVID-19 cases globally may have diluted high-risk factor impacts. Optimized ICU resource management may also explain this discrepancy.

Emerging antifungal resistance in Candida species poses a major public health threat. Resistant *Candida species* correlated with higher mortality rates, such as *Candida parapsilosis* and *Candida auris*, prolonged hospitalization and increased healthcare costs.[25, 26] The burden of candidemia was further exacerbated by the rise of antifungal resistance worldwide. For instance, patients with bloodstream infections caused by resistant Candida species strains harboring the Erg11-Y132F mutation, which confers resistance, exhibited significantly higher crude mortality rates compared to those infected with susceptible strains(63.8% vs. 20%, p=0.008).[25, 27] Additionally, infections caused by fluconazole-resistant Candida parapsilosis had contributed to the increased burden of candidemia. Fluconazole and echinocandins resistance rates reach 20–50% in patients with comorbidities and prior antifungal exposure.[28, 29] While 2016 IDSA guidelines recommended echinocandins, including caspofungin, as first-line treatment for candidemia in both non-neutropenic and neutropenic patients, leading to increased echinocandin utilization.[4] Consequently, echinocandins had become a cornerstone in the treatment of candidemia in recent years. However, resistance to these agents was emerging.[30] Notably, the prevalence of resistant *Candida species* infections exhibited significant geographic variation. Limited access to antifungal agents in Sub-Saharan Africa further exacerbated the global burden of candidemia.[14]

This rising burden is intrinsically linked to increasing antifungal resistance, necessitating enhanced global surveillance systems. Surveillance programs provided valuable data on the prevalence and trends of resistant *Candida species*. Without effective surveillance, the true burden of antifungal resistance may be underestimated, resulting in inappropriate empirical therapy and poorer clinical outcomes. Integrating antifungal stewardship programs with surveillance is critical. Implementation of stewardship programs in tertiary hospitals optimized both cost-efficiency and clinical management of fungal infections while reducing unnecessary empirical and prophylactic antifungal use.[31] It requires healthcare institutions to adopt strategies based on local epidemiology, antifungal resistance patterns, adherence to candidemia treatment guidelines, central venous catheter (CVC) management, ophthalmologic and cardiac evaluations, the best available diagnostic tests, and evidence-based treatment strategies.

## Conclusions

In conclusion, the global burden of candidemia persists, characterized by increasing absolute mortality number despite declining ASMR, reflecting advancements in clinical management. However, significant disparities remained, particularly in resource-limited regions such as Sub-Saharan Africa, where healthcare infrastructure and access to antifungal therapies were inadequate. The emergence of antifungal resistance, notably in *Candida* and fluconazole-resistant *Candida parapsilosis*, further exacerbated the burden, underscoring the urgent need for robust surveillance and antifungal stewardship programs. The COVID-19 pandemic introduced additional challenges, with co-infections significantly elevating mortality risks. Addressing these issues requires targeted interventions for high-risk populations, including males, children, and the elderly, alongside strengthened healthcare systems and improved diagnostic capabilities. Sustained efforts in prevention, early diagnosis, and evidence-based treatment are essential to reduce the global burden of candidemia and mitigate the impact of antifungal resistance.

## Limitations

Although this study utilized robust epidemiological data spanning over three decades to analyze the global burden of candidemia, several limitations warrant acknowledgment. First, the reliance on aggregated data may mask regional heterogeneity. Second, uneven availability of antifungal resistance data across regions may result in underestimation of the contribution of antifungal resistance to mortality. Third, while while addressing COVID-19 impact, the pandemic’s dynamic nature and ongoing effects on healthcare systems may not be fully captured. Finally, mortality projections assume continued advancements in diagnosis and treatment, potentially overlooking unforeseen challenges like emerging resistant strains or healthcare disruptions. These limitations underscore the need for continued research and improved data collection to further refine our understanding of the global burden of candidemia.

## Funding Source

The study was supported by the Anhui Natural Science Foundation (2208085MH195).

## Authorship

All named authors meet the International Committee of Medical Journal Editors (ICMJE) criteria for authorship for this article, take responsibility for the integrity of the work as a whole, and have given their approval for this version to be published.

## Authors’ Contribution

Design of Study and Conceptualization: Lei Zha, Qinghai You. Data Collection: Lei Zha, Hanli Wang, Min Xing, Yonghui Han and Chenghui Wang. Data analysis: Lei Zha, Hanli Wang, Min Xing, Yonghui Han, Haoyu Ji and Shuhui Hu. Original Draft Construction: Lei Zha, Hanli Wang and Ming Xing. Draft Review and Scientific Revisions: Lei Zha, Qinghai You. Final draft approval: all authors.

## Conflict of Interest

The authors have no conflicts of interest to declare.

## Ethical Approval Statement

The study did not require approval because it utilized publicly available data. It adheres to the Guidelines for Accurate and Transparent Health Estimates Reporting for cross-section studies.

## Data Availability

The data utilized in this study were sourced from MICROBE database and publicly accessible datasets. The synthesized data used in this study can be requested from the corresponding authors.

## Acknowledgments

We acknowledge the Institute for Health Metrics and Evaluation (IHME) at the University of Washington for providing access to the MICROBE database. The dataset is licensed for non-commercial use and available at: https://vizhub.healthdata.org/microbe/.

## Notes

### Competing Interest Statement

The authors have declared no competing interest.

### Clinical Trial

NO

